# Supporting young people through the COVID-19 pandemic and beyond: A multi-site qualitative longitudinal study

**DOI:** 10.1101/2023.12.05.23299469

**Authors:** Madelyn Whyte, Emily Nichol, Lisa D. Hawke, Kelli Wuerth, Meaghen Quinlan-Davidson, Aileen O’Reilly, Joseph Duffy, Steve Mathias, JL Henderson, Skye Barbic

## Abstract

Throughout the COVID-19 pandemic, youth have experienced substantial stress due to abrupt changes in education, finances, and social life, compounding pre-existing stressors. With youth (ages 15-26) often at critical points in development, they are vulnerable to long-term mental health challenges brought on by pandemic trauma. To identify youth experiences throughout the pandemic and examine changes over time, we conducted semi-structured interviews among n=141 youth in two Canadian provinces (Ontario and British Columbia) and across the country of Ireland at three time points over the course of more than one year (August 2020-October 2021). We conducted a qualitative longitudinal analysis using an inductive content approach. Categories identified were (1) coping with hardship; (2) opportunities for growth; (3) adapting to new ways of accessing services; (4) mixed views on the pandemic: attitudes, behaviour, and perception of policy response; (5) navigating COVID-19 information; (6) transitioning to life after the pandemic; and (7) youth-led recommendations for government and service response. The findings also reveal trends in health and wellness in accordance with prolonged periods of lockdown, changes in weather, and return to normalcy after the availability of COVID-19 vaccines. Key recommendations from youth include incorporating youth voice into decision making, communicating public health information effectively to youth, enhancing service delivery post-pandemic, and planning for future pandemics. These results provide insights into the extensive longitudinal impacts of the COVID-19 pandemic on young people across three geographical locations. Including youth in decision making for future pandemics or public health emergencies is critical.

## Introduction

Researchers have long studied the relationship between risk and resilience to determine ways to promote positive health outcomes in trauma-exposed youth [1]. Literature exploring the impacts of terrorism and natural disasters has uncovered ripple effects for those exposed to traumatic events in history. Studies on youth mental health symptomology following the 9/11 terrorist attacks revealed links between direct exposure and heightened rates of post-traumatic stress disorder [2–4]. Similarly, data on youth affected by Hurricane Katrina found higher rates of depression and anxiety in the years following the disaster, with secondary stressors of loss of home and financial strain compounding mental turmoil [5,6].

As with discrete traumatic events, the COVID-19 pandemic presented unique challenges for youth (defined here as ages 15-26 years) [7]. Youth reported disruptions in accessing social and recreational services and services for their mental and physical health [8]. For example, while mental health and substance use services quickly pivoted to virtual service delivery modalities, many end users reported not receiving needed services [8,9]. Acute effects of the COVID-19 pandemic on youth include high rates of stress [10–12] and interpersonal challenges [13], gendered differences in mental health status [12,14–17], and mixed effects on substance use [18–20]. Youth also experienced hardships with schooling and academic performance [21–24], with the transition to virtual learning posing challenges for many young people who had difficulties adjusting to a self-directed learning style and lack of supports [24]. Furthermore, the pandemic-related restrictions resulted in increased unemployment rates for youth ages 15-24 in all countries including Canada and Ireland, leading to financial strain and concerns about the future [8,25]. Exposure to heightened stress through sudden risk of infection, school closures, financial strain, isolation, and an increasingly polarized political climate in the early stages of the pandemic created a tumultuous environment with the potential to affect lifetime developmental trajectories [10,11,26].

Quantitative longitudinal studies of youth mental health and wellbeing during the COVID-19 pandemic have suggested ongoing negative impacts of the pandemic as a whole and across many mental health and wellbeing variables [16,17,20,27–29]. Stress, anxiety, and depression increased among youth during the pandemic, while general wellness and positive health behaviors such as physical activity declined [27,28]. However, there is a lack of qualitative, longitudinal literature examining youth experiences during and perceptions of the COVID-19 pandemic and response. An in-depth exploration of youth experiences during the pandemic is required to gain insight into their mental health experiences, perspectives on public health measures, hopes and aspirations for the future, and recommendations for the post-pandemic recovery and future outbreaks, pandemics, and public health crises.

To gain insights into the ongoing experiences of youth during the pandemic internationally and understand how to best meet their needs going forward, we conducted a longitudinal qualitative study during the pandemic. Our objectives were to (1) explore youth experiences during COVID-19 at three time points and in three regions (two provinces in Canada: British Columbia and Ontario, and the country of Ireland) to determine ongoing impacts of the pandemic on mental health and wellbeing, and (2) offer youth-oriented recommendations for government and service response.

## Methods

### Study sample

We recruited participants for this multi-phase longitudinal study from three geographic locations: Ireland and two Canadian provinces [British Columbia (BC) and Ontario (ON)]. Recruitment procedures varied slightly by site as described below. Ethical approval for this study was provided by Jigsaw’s Research Ethics Committee (JREC/2020/004), the Centre for Addiction and Mental Health Research Ethics Board (046-2020), and the University of British Columbia Behavioural Ethics Research Board (H20-01537).

#### Ireland

In Ireland, a mental health seeking sample was recruited from Jigsaw services. Jigsaw – the National Centre for Youth Mental Health is an early intervention integrated mental health service in Ireland that aims to support youth aged 12 to 25 with their mental health and wellbeing [30]. Clinicians invited youth ages 16-25 (with consent from parents/guardians, if under 18) who resided in Ireland and used Jigsaw’s brief intervention services to participate in the study, and, with consent, youths’ contact details were shared with a member of the research team. An invitation to take part in the study was also emailed to all youth registered for Jigsaw’s online synchronous chat support service who had provided consent to be contacted for research purposes. No honorarium was offered to participants.

#### Ontario (ON), Canada

In ON, youth ages 14-28 who were participating in a larger longitudinal quantitative study on mental health and COVID-19 at the Centre of Mental Health and Addictions (CAMH) [31] were asked for consent to be contacted for this sub-study. CAMH is the largest mental health teaching hospital in Canada, providing a range of clinical services for all ages. The study included youth recruited from three clinical studies and one non-clinical study [32–34]. Among those who provided consent, emails were sent to potential participants, and purposive sampling was carried out to include a diverse sample of youth in this sub-study. Thirty participants were recruited from the clinical sample, and thirty-one participants were recruited from the non-clinical sample, with the goal of representing both cohorts and a wide range of demographic characteristics. Participants received an email message inviting them to participate. Participants were paid a $30 honorarium for each interview.

#### British Columbia (BC), Canada

In BC, a Foundry-led social media campaign was developed with youth and launched to recruit diverse youth ages 16-24 who resided in BC. Foundry is an integrated youth services initiative for youth ages 12-24 that provides support in person and virtually through a single access point for multiple service streams, including mental health [35]. Among those who indicated interest in the study, purposive sampling was carried out, and a diverse sample of youth from across the province was invited to the consent phase of the study. Two groups were recruited: A clinical group (n=30) who had accessed Foundry services in the past 12 months, and a non-clinical group (n=30) who had not accessed Foundry services. As with ON, participants in BC were paid a $30 honorarium for each interview.

### Consent and interview procedures

Data collection procedures at each site varied slightly. Youth provided verbal or written informed assent/consent to their data being collected and stored for research purposes. In cases where youth could not legally consent due to jurisdictional laws, consent was received from their parent/guardian. Participants were informed that all questions were optional, and that they could end the interview at any time. Participants were followed up with via email. The interview schedule is reported in Table 1.

**Table 1.** Interview schedule across all sites.

|  | **T1** | **T2** | **T3** |
| --- | --- | --- | --- |
| **Ireland** | November 2020 –<br>January 2021 | February – March<br>2021 | August 2021 |
| **Ontario** | August 2020 | March – April 2021 | September – October<br>2021 |
| **British Columbia** | November 2020 –<br>January 2021 | March – June 2021 | August – October<br>2021 |

In ON, demographic data (e.g., age, gender identity, race/ethnicity, employment status) and psychosocial data were collected from the initial quantitative survey [36]. Interviews were conducted by phone or through a secure video conferencing system, WebEx (Cisco Systems, San Jose, California), hosted on a secure institutional server. Interviews were digitally recorded and transcribed verbatim by either a research staff member or a professional transcription agency. Participants were assigned unique participant IDs, and identifying information was removed during transcription. In Ireland and BC, demographic surveys were completed electronically at the start of all three time points. In BC, psychosocial questions were completed as part of this survey. Semi-structured interviews were conducted through Zoom (Zoom Video Corporation, San Jose, California) by a member of the research team. In BC, this research team for interviews consisted of a research coordinator and seven youth research assistants, and in Ireland the team consisted of two youth research assistants. Interviews were recorded and transcribed verbatim by two youth research assistants in BC and two youth research assistants in Ireland. Unique participant IDs were given and identifying information was removed from the transcripts to maintain confidentiality. A copy of the semi-structured interview guides used across all sites is contained in Appendix A.

The engagement of young people was a core component of this study. Youth co-researchers were employed at each site to support all elements of the research process and were provided with training and supervision by the lead researchers. In addition, youth reference and co-researcher groups were consulted on various aspects of the project across the three study sites, including co-developing the recruitment materials and interview questions, interpreting and validating the findings, and contributing to other knowledge translation materials [37,38].

### Analysis

We employed inductive content analysis, which involved reading and re-reading the data (familiarisation); identifying broad categories; developing and refining subcategories and fine-grained codes; and collating and interpreting the data to generate succinct categories and subcategories [39]. NVivo 12 Software [40] was used to support data analysis. This process was iterative, with many drafts of refinements until an accurate depiction of the data was captured [39]. At the intra-site level, the research team met weekly to ensure a mutual understanding of code definitions and coded two transcripts together for each time point. At the inter-site level, all team members met weekly to discuss discrepancies and refine codes and the coding framework.

## Results

### Demographics

As shown in Table 2, a total of 141 participants ages 14-26 years took part in the study at T1 (BC n=59, ON n=61, Ireland n=21). Retention was 87% at T2 (n=123 total; BC n=50, ON n=56, Ireland n=17), and 78% at T3 (n=110 total; BC n=43, ON n=53, Ireland n=14).

**Table 2.** Sociodemographic characteristics of participants included in the study at T1.

|  | BC (n=59) | ON (n=61) | Ireland (n=21) | Total (n=141) |
| --- | --- | --- | --- | --- |
| **Age (years)** |  |  |  |  |
| 14-17 | 12 (20.3%) | 4 (6.6%) | 7 (33.3%) | 23 (16.3%) |
| 18-21 | 27 (45.8%) | 30 (49.2%) | 10 (47.6%) | 67 (47.5%) |
| 22-25 | 20 (33.9%) | 26 (42.6%) | 4 (19.1%) | 50 (35.5%) |
| 26 | 0 (0%) | 1 (1.6%) | 0 (0%) | 1 (0.7%) |
| **Gender** |  |  |  |  |
| Boys/Men | 12 (20.3%) | 22 (36.1%) | 2 (9.5%) | 36 (25.5%) |
| Girls/Women | 43 (72.9%) | 29 (47.5%) | 18 (85.7%) | 90 (63.8%) |
| Non-binary | 0 (0%) | 6 (9.8%) | 1 (4.8%) | 7 (5.0%) |
| Transgender | 1 (1.7%) | 1 (1.6%) | 0 (0%) | 2 (1.4%) |
| Another gender identity | 3 (5.1%) | 3 (4.9%) | 0 (0%) | 6 (4.3%) |
| **Race/Ethnicity** |  |  |  |  |
| Caucasian | 25 (42.4%) | 24 (39.3%) | 17 (81.0%) | 66 (46.8%) |
| Black | 0 (0%) | 6 (9.8%) | 2 (9.5%) | 8 (5.7%) |
| Asian | 18 (30.5%) | 20 (32.8%) | 2 (9.5%) | 40 (28.4%) |
| Latin American | 1 (1.70%) | 1 (1.6%) | 0 (0.0%) | 2 (1.4%) |
| Mixed heritage | 8 (13.6%) | 8 (13.1%) | 0 (0.0%) | 16 (11.4%) |
| Ashkenazi Jewish | 0 (0.0%) | 1 (1.6%) | 0 (0.0%) | 1 (0.7%) |
| Middle Eastern/North African | 5 (8.5%) | 0 (0.0%) | 0 (0.0%) | 5 (3.6%) |
| Métis | 1 (1.7%) | 0 (0.0%) | 0 (0.0%) | 1 (0.7%) |
| Missing | 1 (1.7%) | 1 (1.7%) | 0 (0.0%) | 2 (1.4%) |

Table 3 outlines psychosocial factors documented across time-points in BC and ON. Due to resource limitations, participants from Ireland did not complete the psychosocial surveys.

**Table 3.** Psychosocial characteristics across timepoints in BC and ON.*

|  | BC |  |  | ON |  |  |
| --- | --- | --- | --- | --- | --- | --- |
| Employment Status | T1 | T2 | T3 | T1 | T2 | T3 |
|  | (n=59) | (n=41) | (n=27) | (n=60) | (n=54) | (n=51) |
| Employed | 10<br>(17.0%) | 9<br>(22.0%) | 7<br>(25.9%) | 6<br>(10.0%) | 14<br>(25.9%) | 16<br>(31.4%) |
| In school | 24<br>(40.7%) | 14<br>(34.1%) | 4<br>(14.8%) | 35<br>(58.3%) | 16<br>(29.6%) | 12<br>(23.5%) |
| Both | 23<br>(39.0%) | 16<br>(39.0%) | 12<br>(44.4%) | 9<br>(15.0%) | 19<br>(45.2%) | 20<br>(39.2%) |
| Neither | 2 (3.4%) | 2 (4.9%) | 4<br>(14.8%) | 10<br>(16.7%) | 5 (9.3%) | 3 (5.9%) |
| Physical Health | T1 | T2 | T3 | T1 | T2 | T3 |
|  | (n=59) | (n=41) | (n=27) | (n=59) | (n=54) | (n=51) |
| Poor to Fair | 9<br>(15.3%) | 8<br>(19.5%) | 3<br>(11.1%) | 19<br>(32.2%) | 20<br>(37.0%) | 18<br>(35.3%) |
| Good to Excellent | 50<br>(84.8%) | 33<br>(80.5%) | 24<br>(88.9%) | 40<br>(67.8%) | 34<br>(62.7%) | 33<br>(64.7%) |
| Mental/Emotional Health | T1 | T2 | T3 | T1 | T2 | T3 |
|  | (n=59) | (n=41) | (n=27) | (n=59) | (n=55) | (n=51) |
| Poor to Fair | 33<br>(55.9%) | 19<br>(46.3%) | 10<br>(37.0%) | 28<br>(47.5%) | 23<br>(41.8%) | 22<br>(43.1%) |
| **Good to Excellent** | 26 | 22 | 17 | 31 | 32 | 29 |
|  | (44.1%) | (53.7%) | (63.0%) | (52.5%) | (58.2%) | (56.9%) |

\* Some participants did not fill in demographics at all time points and some surveys were partially complete.

### Content categories

Table 4 outlines seven primary categories and multiple subcategories uncovered during analysis. Interviews at different time points and sites were analyzed separately and trends over time and across sites were documented, capturing shifts in health and wellness, personal circumstance, and perception of policy response. These findings are discussed together when consistent across sites and time points, and differences are described when applicable. These categories and subcategories are described in the following sections and include verbatim supporting quotes.

**Table 4.**
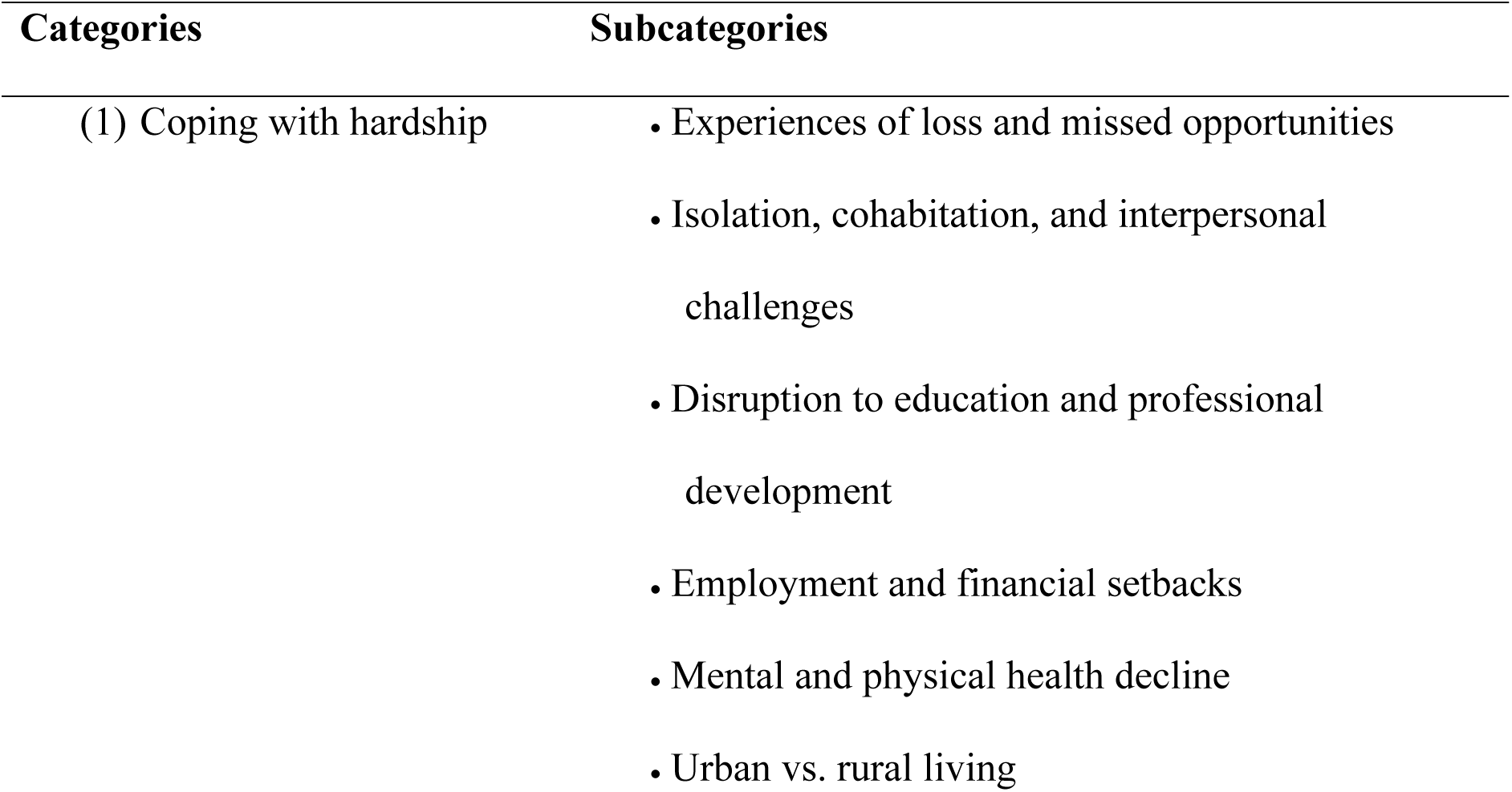
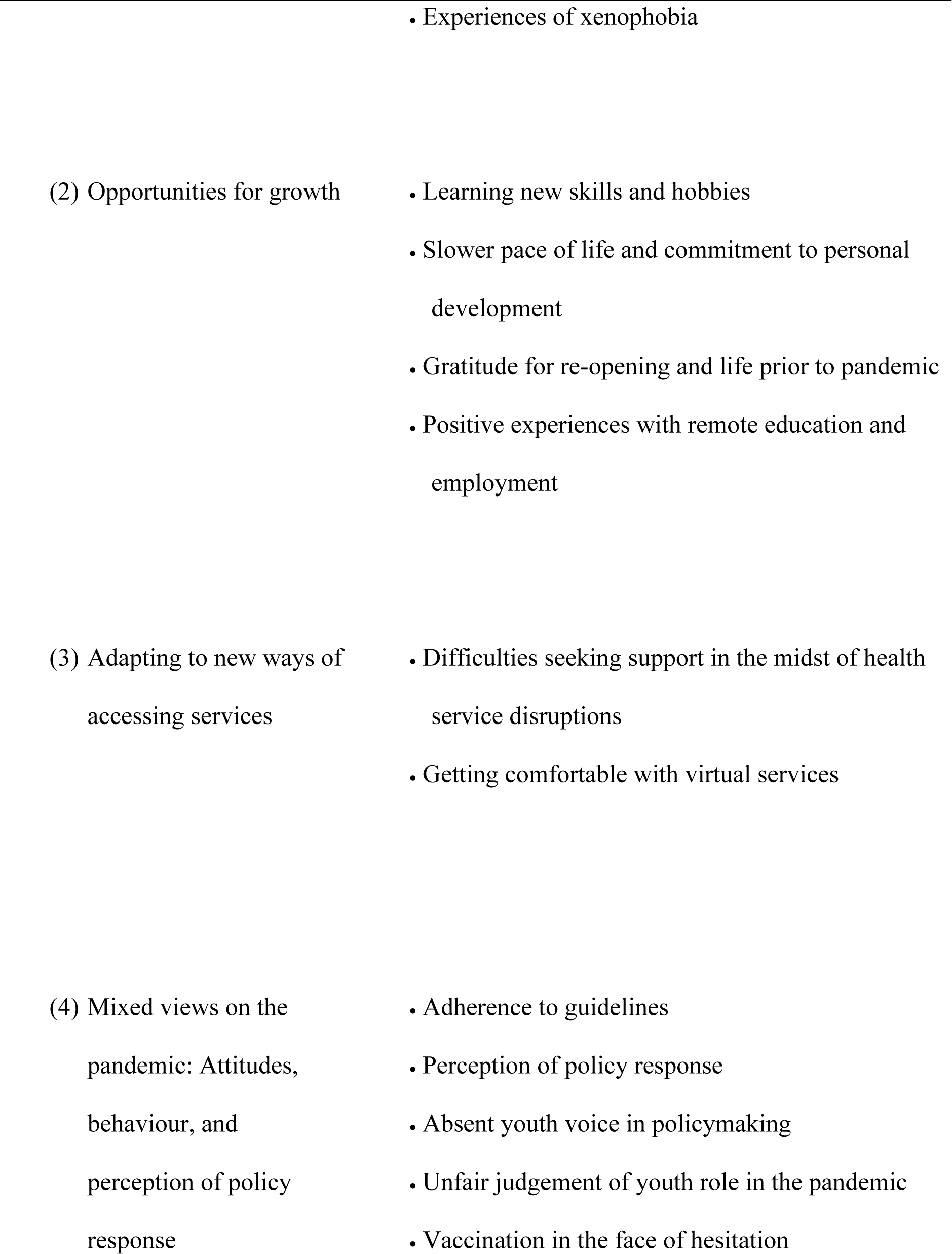
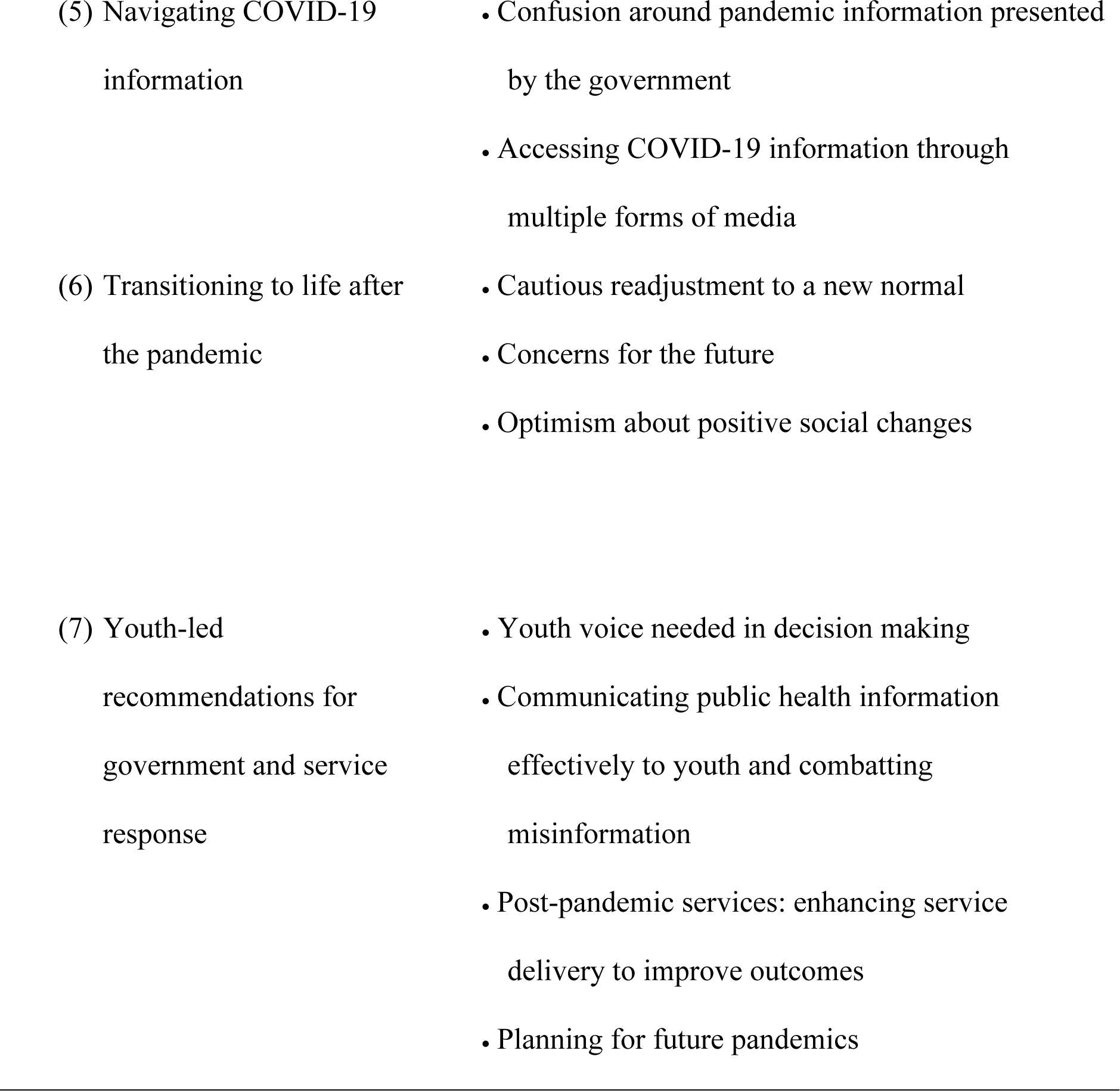
Categories and subcategories.

| Categories | Subcategories |
| --- | --- |
| (1) Coping with hardship | • Experiences of loss and missed opportunities<br>• Isolation, cohabitation, and interpersonal challenges<br>• Disruption to education and professional development<br>• Employment and financial setbacks<br>• Mental and physical health decline<br>• Urban vs. rural living |
|  | • Experiences of xenophobia |
| (2) Opportunities for growth | • Learning new skills and hobbies<br>• Slower pace of life and commitment to personal development<br>• Gratitude for re-opening and life prior to pandemic<br>• Positive experiences with remote education and employment |
| (3) Adapting to new ways of accessing services | • Difficulties seeking support in the midst of health service disruptions<br>• Getting comfortable with virtual services |
| (4) Mixed views on the pandemic: Attitudes, behaviour, and perception of policy response | • Adherence to guidelines<br>• Perception of policy response<br>• Absent youth voice in policymaking<br>• Unfair judgement of youth role in the pandemic<br>• Vaccination in the face of hesitation |
| (5) Navigating COVID-19 information | • Confusion around pandemic information presented by the government<br>• Accessing COVID-19 information through multiple forms of media |
| (6) Transitioning to life after the pandemic | • Cautious readjustment to a new normal<br>• Concerns for the future<br>• Optimism about positive social changes |
| (7) Youth-led recommendations for government and service response | • Youth voice needed in decision making<br>• Communicating public health information effectively to youth and combatting misinformation<br>• Post-pandemic services: enhancing service delivery to improve outcomes<br>• Planning for future pandemics |

### 1. Coping with hardship

Several pandemic-related hardships were identified, amounting to cumulative stress over the course of the study period. Participants detailed challenges pertaining to loss of experiences, interpersonal relationships, disruption to education and professional development, financial setbacks, mental health decline, and experiences of xenophobia exacerbated by a rise in political tension.

#### Experiences of loss and missed opportunities

Youth reported experiencing loss of important childhood and adolescent milestones, describing missing out on their *“prime years”* (BC, T1) due to lockdowns and restrictions. Youth expressed awareness for how their generation has been impacted in unique ways due to the transitional nature of the participants’ ages, with one participant saying, *“I’ve been a lot more lonely … it feels like a ton of my prime-time – my prime years are being wasted”* (BC, T1). For some, these disruptions felt like a disadvantage in terms of emotional and professional development, with one participant saying, *“It affects our generation a lot, just because … these are our prime years to meet new people and gain new experiences and stuff like that. I feel like we have … a disadvantage”* (BC, T1).

Youth were also impacted by missing out on important events to celebrate achievements such as graduation, birthdays, and other moments of significance. These missed opportunities to recognize pivotal moments of accomplishment in a public forum were realized in the early days of the pandemic: *“I didn’t have a graduation ceremony, so I didn’t get to celebrate or take pictures with my parents. Didn’t get to walk with my diploma or say goodbye to my friends, or do any form of partying or celebrating finishing school”* (BC, T1).

No longer able to engage in typical activities as an adolescent, youth suffered erasure of formative experiences, reporting, *“you’re never going to get the year back”* (Ireland, T2). Lost time and missed opportunities contributed to a collective sense of grief.

#### Isolation, cohabitation, and interpersonal challenges

Across regions, interpersonal challenges became apparent due to lockdown and restrictions on social gatherings, where connections became strained due to diminished contact: *“It is almost like a whole year of bonding is gone”* (Ireland, T1). Warmer weather acted as a buffer against isolation, since participants had more freedom to engage with others safely outside. During colder months, as case numbers rose and restrictions tightened, participants expressed increased feelings of loneliness:

> “*The summer, at least in BC, was still pretty good in terms of being able to see people … The weather being good, as well, helped. Since then, it’s been tough not seeing people in person, and not having those kinds of social connections”* (BC, T2).

Different views on policy guidelines and pandemic restrictions contributed to relational disconnect. The strain on relationships was most apparent at T2 as restrictions tightened, and friends and family members had differing views on vaccinations and following guidelines:

> “*I still feel kind of disconnected from a lot of my friends, just because of the pandemic, and everyone’s different ways that they feel about the restrictions. A lot of my friends are, maybe, more open, and maybe not following the rules as much as myself”* (BC, T2).

Many participants expressed technology fatigue over time, where communicating in the form of texting or video chatting could not replace the in-person connection: *“That’s a big part of it, not being able to just hug people”* (Ireland, T2). For some, maintaining virtual relationships was a daunting task, described more like a chore than an opportunity for social interaction: “*You kind of can only keep so many relationships going virtually”* (ON, T2). As one participant described the trials and tribulations of virtual communication:

> *“I just have a lot of troubles with calls, maybe not calls, but texting people … I just don’t think you can maintain any sort of social relationship just on social media.*” (ON, T1).

In contrast, too much time with family enabled conflict due to a lack of personal space, with youth noting the adverse effects on their mental health as a result. *“Family issues spiked up”* (Ireland, T1) during lockdown, when family members were living, working, and attending school under one roof, the effects of which were felt most among youth with challenging dynamics with their parents: *“My parents weren’t very supportive of me and I kind of had to stay in that house, and you know, get talked down to and belittled every single day, but also my anxiety from not going out really anywhere skyrocketed”* (ON, T1). Confined by the inability to seek refuge in safe relationships, feelings of isolation were compounded for youth in challenging family dynamics.

#### Disruption to education and professional development

Youth described how online learning was an adjustment. Difficulties associated with abrupt changes in educational formats were most apparent at T1. Youth experienced obstacles managing a self-directed learning style, feeling distracted and less productive at home. Some students with disabilities experienced challenges navigating a new way of learning without the necessary infrastructure to provide accommodations in virtual course formats:

> *“A lot of institutions took the opportunity of digital and online services to mean that they no longer had to provide specific disability services, which, had they talked to students, would know that’s not acceptable nor equitable access … online does not mean accessible”* (BC, T1).

Technical issues compounded by adjusting to an online format while at home posed several learning barriers:

> *“I find it very hard it’s like school online takes a huge toll on like me personally I find it extremely stressful to keep up with it. It’s given me a couple of like breakdowns especially in the last couple of weeks … it doesn’t feel like you’re actually in school like or in college. You’re just in a room watching things”* (Ireland, T1).

For those experiencing greater mental health problems due to pandemic-related challenges, academic performance declined*: “It really affected me, and it really affected my mood. I couldn’t do well on online school, so I really was in trouble, and then I didn’t finish”* (BC, T1). Students described heightened stress, lower mood, lack of focus, an inability to complete course work, and an overall work environment not conducive to productivity.

Teacher-student communication issues emerged, where students needing extra assistance were unable to receive proper support virtually. Managing questions over email was not adequate for those requiring clarity and a fluid back-and-forth dialogue on an academic issue:

> *“We used to speak with the teacher, we used to be in a classroom, you could talk to them. But now everything’s online, and learning online is a lot harder, ‘cause you can’t really ask questions the way you could in class sitting in front of the teacher”* (ON, T1).

In applied programs and career paths requiring more technical training among college- and university-aged students, educational progress slowed. Without adequate hands-on experience, youth felt they were not receiving the same type of education they would have had if the pandemic had not happened. In some cases, youth worried how lacking such skills would affect future career prospects, feeling *“ripped off in terms of professional development”* (BC, T2).

> *“Starting a brand-new program, especially one that’s super hands-on learning, it was really difficult to find times and opportunities to practice hands-on skills that I need to develop”* (BC, T2).

#### Employment and financial setbacks

Job loss or reduced *“COVID hours”* (BC, T2) led to financial stress that was particularly noted in T1. Fear surrounding being able to find a new job and afford the cost of living took a toll emotionally, adding new stress for some on relationships:

> *“It was the end of my school term, so I had no money left, and it was definitely pretty scary, so I don’t want to be in that situation where I was floundering, looking for money, and I ended up having to rely on my boyfriend a lot. Which, this is my ex-boyfriend, but it kind of created this weird power imbalance in our relationship, where I had to rely on him for everything”* (BC, T1).

As an age group that occupies a large portion of customer service and hospitality positions, youth expressed concern about returning to a work environment with potentially increased exposure to COVID-19: *“[Working in retail is] absolutely terrifying, to be exposed to that many people every day, like, you know, how many people come through to shop which is something that we’ve all been avoiding over the past couple months”* (Ireland, T1). Fear of contracting COVID-19 arose, along with challenges dealing with the public with the implementation of new public health mandates requiring masks and vaccination in certain establishments:

> *“Since everything was opening up, the public seemed a little more lax in terms of some of the public health restrictions. So, you know getting into disagreements over, you know if they should wear a mask and stuff like that, and having to explain that the pandemic is still going on. It is a customer service job, so I’d get yelled at quite a lot this summer* [2021] *over that, as well as you know trying to enforce public health recommendations”* (ON, T3).

#### Mental and physical health decline

Mental health challenges presented in the form of disrupted sleep schedules, increased substance use and social anxiety, and decreased motivation. Youth described how prior stressors were exacerbated by pandemic-related stress, with symptoms most prevalent during lockdown periods and winter months.

With online school and reduced work hours or job loss, maintaining a routine became difficult, which perpetuated mental health challenges: *“I have trouble with maintaining a regular sleep schedule. And so that’s exacerbated by not having a schedule – like during the pandemic there’s more onus on me to schedule things and there’s less outside – outside factors for me to rely on as a schedule”* (ON, T2). Youth described reliance on unhealthy coping mechanisms and increased substance use to mitigate mental health challenges: *“When I feel like that [out of control], then that’s when I start to smoke and do a lot of other things to keep myself distracted and mellow it out”* (ON, T1). Unable to turn to typical coping strategies such as leaning on friends, youth struggled to find ways to support their wellbeing:

> *“It’s been significantly more difficult to manage symptoms since the COVID pandemic has hit, and I think that’s because, not being able to get out of the house and see my friends … socializing, getting out, and doing things, is quite important to me – keeping a good mental health”* (BC, T1).
>
> *“My substance use has definitely gone up or it has become more frequent, like quite dramatically. And that’s in part because I, again, I live alone, I work from home, so there’s a lot more opportunity sort of for me to engage with substance use. And also because I’m engaging with that more and I have more time to sort of think to myself and ruminate on my life circumstances and my situation and stuff like that, that has also been challenging to my mental health in general which has also contributed to me using substances more and more”* (ON, T2).

Over time, youth described how isolation heightened social anxiety: *“You’re always on edge, everything seems like we’re in war or something and you have to stay inside ‘cause there’s curfews and danger and everything all the time”* (Ireland, T2). Living in a perpetual state of fear when interacting with others and experiencing lockdowns led to difficulties socializing: *“Being cooped up inside 24/7 isn’t good for anybody … You stay inside, you’re going to develop different problems socially”* (Ireland, T2). Everyday tasks such as going to the grocery store or seeing friends became associated with fear:

> *“I’m afraid of going into public places like Costco, or being in a bigger group of friends. If there’s ten people – even if we’re social distancing, I still feel uncomfortable if someone’s gone somewhere, and I know where they’ve gone, or I don’t know where they’ve gone even, I feel uncomfortable”* (BC, T2).

Motivation to learn, work, or complete typical tasks decreased while navigating life through the mundaneness of the pandemic: *“I can’t bring myself to do anything … If I tried to do school, I couldn’t do school, even one course. I can barely take care of myself throughout the day. I just don’t have any motivation to do anything”* (ON, T3).

For many, winter months intensified boredom, where shorter days and colder weather negatively impacted mental health. Youth described feeling like they were living the same day on repeat:

> *“It definitely got worse in the winter. God, it’s much harder to do anything when it’s cold. I don’t know, especially I think in January* [2021] *was terrible, like when it was getting dark, ugh, I hated that month. And it was kind of, there was nothing else to do but just wake up. I kind of started to feel like I was living the same day over and over again, especially in, like January, February, like I was living the same week because it would be like I have the exact same worries on Monday and then that’s done. And then I have the exact same worries again and again and again and again”* (Ireland, T2).

In instances where youth contextualized prior difficulties with mental health problems, COVID-19 added *“this extra layer of anxiety and stress that’s just been coated across everythin … It feels like it’s just blanketed the whole thing, and it’s added more weight to everything”* (BC, T1). Youth described feeling a *“constant high state of alert and stress”* (BC, T2), with little to no relief: *“It’s just something that’s always there and I feel like it kind of accentuates whenever something that you do have that’s more negative on your life … It kind of compounds with anything negative in your life, and so it’s very easy to kind of wear down on you”* (ON, T2).

In addition to negative mental health impacts of the pandemic, youth experienced physical health declines due to closure of gyms and recreation centres, colder weather making it difficult to exercise outdoors, and inaccessible physical health services, including doctors, physiotherapy, and other health services: *“I loved going to the gym but I have not been to a gym since probably December”* (ON, T2).

However, improved mental and physical wellbeing were often observed in summer months, where youth had opportunities to get outside, exercise, and safely interact with peers in a physically distanced manner: *“I think summer is also a big factor, like weather. It changes everyone’s mood. So I think it’s definitely more hopeful when the summer comes. Everyone will be more happier and more hopeful”* (BC, T2).

#### Urban vs. rural living

COVID-19 experiences differed for youth depending on their community. Those in rural areas expressed gratitude for more open space to get outside and physically distance:

> *“If I was to live in town in the city, I wouldn’t be going out for a walk, because I’d be more likely to bump into people … I’d be inside more, and then I’d be driven more crazy. I really do need those walks every now and again, and just get out of the house. And I don’t know if I would be confident enough to do that in town”* (Ireland, T2).

However, misinformation and people not following guidelines was a reported concern, with one youth in a rural community reporting, *“I think in terms of the general attitude around COVID, it’s maybe a bit more … Uneducated, than in a city”* (BC, T2). In urban spaces, greater population density led to an increase in fear and added challenges getting around the city: *“If we had enough money to afford a car, we wouldn’t have to worry about catching COVID from public transit. Wouldn’t have to get in that sardine bus. We’d be able to go and visit friends without worrying about, like, OK, I am exposing myself to, like, hundreds of people”* (ON, T3).

#### Experiences of xenophobia

Asian-Canadians experienced a *“big increase in harassment and racism”* (BC, T2) noted midway through the study period. Instances of hateful rhetoric became more commonplace, and as a BC participant recalled, *“I did get some of the racist comments when I was at Costco, because of being Asian. I did receive a lot of comments about it, so I think those did affect me, and I think it’s going to affect me after the pandemic, as well”* (BC, T2). Because of these experiences, as well as seeing cases of racism and xenophobia on the news, some described feeling unsafe in public settings:

> *“I guess it’s important to stay more vigilant now. (…) [S]ometimes there’s those kind of creepy looking [people] who you don’t know if they’re going to start verbally harassing you on the subway and stuff because you’re alone and there’s not people around to kind of protect you anymore”* (ON, T3).

In Ireland, one participant spoke about how xenophobia is resulting in a “*divide in society*”, citing a mentality in Irish society of *“‘No Chinese, no Brazilians, no Brits’, because like they brought the new variants”* (Ireland, T2). They also spoke about their family members experiences’ *“getting abused on the bus”* and being told “*you brought COVID here*” (Ireland, T2).

### 2. Opportunities for growth

#### Learning new skills and hobbies

The pandemic presented an opportunity to develop new hobbies and skills. Honing a craft, indulging in creative endeavors, and exercising buffered against stressors in the early days of quarantine during T1 interviews:

> *“During the first lockdown, I actually picked up the hobby of running, because I didn’t have anything else to do. And I always wanted to try running, but I never really got the chance to”* (Ireland, T1).
>
> *“I started doing baking as a hobby more during COVID. I’ve always liked baking, but I really picked it up a lot more throughout COVID”* (Ontario, T1).

With newfound leisure time, youth were able to put their energy towards activities that would otherwise not be prioritized, which improved mental health:

> *“Especially during the early days of quarantine, that was pretty helpful, being able to create something, or lots of people got into baking. I don’t know, I think little things like that, like being able to acquire a new skill that you didn’t have before, I think that’s important for your mental health, and just feeling like you are accomplishing something, even when everything else is kind of at a standstill”* (BC, T1).

#### Slower pace of life and commitment to personal development

Many used their newfound free time to slow down, self-reflect, and *“build better habits”* (ON, T3). The impacts of a slower pace of life were particularly apparent during T1 interviews, as some youth experienced reduced responsibilities because of changes in employment and education. This gave youth time to reflect: *“It’s forced me to slow down, in a way, and to spend more time with myself, and learn more about myself”* (BC, T1). Others were able to recover from burnout: *“I was running at such a high level, and burning a lot of steam and not giving myself that time to reflect on situations and myself, and COVID kind of forced me to do that, because I had the literal time to do it”* (BC, T1).

Despite hardship during the pandemic, more time alone acted as a catalyst for change: *“It taught me a lot about my mental health … how to take care of myself, and how to practice self-care”* (BC, T1). Several youth took initiative to understand themselves better and begin a journey of healing during this time.

In some instances, youth noted an increase in help-seeking behaviour, declaring, *“If it wasn’t for the pandemic, I think I’d still probably be suffering from mental health [problems]”* (Ireland, T3). These participants practiced self-care by confronting their mental health issues and reaching out for support instead of suppressing emotions:

> *“The thing about being stuck in your house is that you can’t escape anything. Like all of your issues, you have to deal with them right now because there’s no going anywhere. Like it’s just you and your head in this house so you have to, you know, deal with it … I’m at a point now where I’m being forced to make life changes and I’m being forced to actively confront my life and my issues”* (Ireland, T2).

#### Gratitude for re-opening and life prior to the pandemic

Gratitude and appreciation toward returning to a “normal” life as restrictions lifted were noted across all time points: *“I think it kind of just made me appreciate everything, kind of more. Like even whenever I was able to see my granny for the first time, I just would be something so normal to me and then it was just like this whole big thing”* (Ireland, T1). For some, the pandemic served as a reminder to *“[have] gratitude for what’s happening in the present cause you don’t really know what’s going to happen in the future”* (ON, T1).

#### Positive experiences with remote education and employment

While some youth experienced a multitude of difficulties adjusting to remote education and employment, others found they were able to excel with the shift to virtual. Positive attitudes towards remote education and employment increased in T2 and T3 as rocky transitions passed. Virtual education and employment were valued for accessibility reasons for individuals who live in rural regions and/or have certain disabilities or physical or mental health conditions that limit their ability to participate in in-person activities: *“I also have a chronic illness, which affects fatigue and mobility, so being able to have class online was really helpful, as well”* (BC, T1).

### 3. Adapting to new ways of accessing services

Physical health, mental health, and substance use services had to quickly shift from in-person to virtual care. During this transition period, youth experienced service disruptions, noted in T1 interviews. Barriers to accessing services improved with time as adaptations to online service delivery developed. However, concerns including prolonged wait times, not feeling prioritized in the mental health sector, and hesitancy to seek help persisted through all time points.

#### Difficulties seeking support in the midst of health service disruptions

For many, wait times were an obstacle prior to the COVID-19 pandemic, made worse by service closures during lockdowns. Such disruptions left youth with few resources to manage their mental health: *“There was no options. There was no Skype call offered to me. There was literally nothing. I was lucky to receive a text message at one stage”* (Ireland, T2).

With nowhere to turn, several youth noticed a decline in health:

> *“For myself, though … Maybe some worsened health, just because appointments have taken so long to get to, and I haven’t been able to see in-person doctors”* (BC, T1).

#### Getting comfortable with virtual services

Once virtual services were implemented, some youth expressed preference for in-person support, though many acknowledged benefits of virtual appointments. One was using virtual services for general health concerns and prescription refills:

> *“It’s been working well, for me. Most of the time, it’s just for a prescription refill or an update on how my health has been, so it’s pretty convenient to just call and expect a phone call, like I can be at home, and I don’t have to go anywhere and wait for too long”* (BC, T1).

Another benefit was that many participants felt more comfortable accessing virtual supports than in-person supports due to the online disinhibition effect:

> *“When I was doing it face to face I would shy away from a lot of conversations because I’d feel embarrassed or I’d get distracted. Where now on a Zoom call it just kind of feels like a normal phone call. Ahm, so I’d kind of be able to tell a lot more because I can’t find myself distracted because all I can see is obviously my screen”* (Ireland, T1).

By T3, the convenience and accessibility aspects of virtual services were acknowledged as something that should be continued: *“I think online services can be great, more accessibility for people in remote locations. Jobs that don’t conform to like business hours for therapists … I think it’s better than only in person, because in person is just not accessible”* (BC, T3).

On the other hand, some participants had ongoing negative experiences with virtual services. The effects of their day-to-day life being virtual began to take a toll: *“I’m so fatigued from all the virtual stuff that it would just drive me crazy if I tried to use virtual services for mental health”* (ON, T3).

Technology problems created fractures in the flow of appointments, negatively impacting the overall quality and experience of services:

> *“When the network is cut off or the internet’s lagging or you’re having a hard time hearing the other person, that interrupts the intervention. That interrupts the mindfulness practice that you were just in the middle of and kind of taints the experience”* (ON, T3).

Disrupted continuity impacted client satisfaction, reducing desire to access services in this way. Participants described virtual counselling as *“totally disconnected”* (BC, T1). Lack of a personal connection to establish a rapport contributed to hesitation attending more intimate appointments over phone or video*: “I feel discouraged to go up to these online sessions. It still feels like you’re talking to the computer. It’s not the same”* (Ireland, T1).

For youth living with family or roommates, they felt the lack of privacy led them to be hesitant to access virtual services and inhibited from maximizing their therapeutic experience:

> *“I wasn’t sure about [virtual services]… if I was able to do that in the home, because as I said before, like, everybody’s at home. It’s hard to find a space where it’s quiet and nobody can hear you. A space where you can really be vulnerable with the person that you’re talking to and trust that it’s only them that can hear the information and all that”* (Ontario, T1).

With regards to synchronous chat, the depersonalization aspect of texting was a concern due to inability to convey the full range of expression during diagnostic procedures: *“A lot of emotions and undertones aren’t really gotten across with text … from a diagnostic perspective, it’s probably not the best because people can really fake things via text, like, you could lie your way out of anything on text message, and nobody would know”* (ON, T1). For others, synchronous chat provided an alternative option when coping with anxiety due to flexibility of communicating in times of need without a scheduled appointment and no pressure: *“It’s not invasive or anything, and you can say as much as you want or stop replying when you want”* (Ireland, T1).

### 4. Mixed views on the pandemic: Attitudes, behaviour, and perception of policy response

#### Adherence to guidelines

Notable observations were made regarding how youth and other generations reacted to the pandemic. Fear of the unknown meant a greater willingness to follow public health guidelines such as physical distancing, wearing a mask, and frequently using hand sanitizer in the early days of the pandemic:

> *“I think initially everyone was [taking it seriously], young and old, were petrified, when we didn’t know. I think fear of the unknown at the start was why the first lockdown worked so well. Because everyone was really adhering, because like, no one knew how it [COVID-19] was going to affect you, you’d see the awful things on the TV and you didn’t know that sometimes you could literally have nothing [no symptoms]”* (Ireland, T1).

Others expressed willingness to adhere to public health guidelines in service of collective responsibility: *“I think that their role, like our role, everybody’s role is to try and stop the spread. So everybody has a responsibility to limit their contact”* (Ireland, T1).

Several youth expressed discontent regarding others not taking guidelines seriously, impacting how youth perceived the longevity of the pandemic: *“I also do see people around me that are not taking the pandemic very seriously … I just have frustrations with the current situation that make me not very optimistic about it”* (BC, T1). Social media became a space where youth could check in on what their peers were doing, influencing feelings of hope during this time: *“I see a lot of Instagram posts, social media posts where people are all together without a mask on, without anything, large group of people. That definitely brings down the hope a little bit … Situations like that can really set back the progress that’s been made”* (ON, T1).

Youth perceived their peer groups as adhering to guidelines more than older generations in terms of physical distancing: “*I feel like it’s usually the middle-aged people who are more not following the policies and rules of outdoor places, or even restaurants and whatnot. I think that for the most part, youth are being pretty respectful of that”* (BC, T1). Potential reasons for this difference were explored:

> *“I think that teenagers as a whole have actually handled it very, very well. Because we’re so young, we’re like a sponge. We soak things up. We’re still being taught things. We’re still being told how to do things, as where, somebody in their 30s are kind of set in their ways”* (Ireland, T1).

Over time, youth expressed less regard for following public health guidelines: *“The fedupness has taken over the wanting to be safe”* (Ireland, T3). Participants discussed growing tired of repeated lockdowns and constantly changing guidelines: *“Pandemic fatigue has people saying ‘Well whatever, it hasn’t touched me, yet. Why would it touch me, now?*’ *And I feel like people are kind of over all the rules and regulations”* (BC, T2). As people became accustomed to a new way of living in the midst of the COVID-19 pandemic, there was a reduced sense of urgency to follow restrictions:

> *“I think, in the beginning we all saw how serious it was and we all kind of knew it was our, not job, but aim, to keep other people safe, so that’s why we were sticking to all the guidelines and we were doing what we were told and things like that. But in the past three months, being stuck at home, back lockdown again, school. I’ve seen a lot of people, and it shocked me, that have just completely given up on it, and just don’t want to do it anymore … I think they’ve kind of given up hope a little bit”* (Ireland, T2).

In this sense, youth described this change in behaviour as not a result of lack of caring, but rather a progression of the impact of the pandemic on lifestyle change and mental health. This increasing toll contributed to a sense of indifference towards following rules. Despite the disillusionment with policy guidelines, increased anxiety about getting COVID-19 was also noted over time:

> *“I think I’m more actively worried than I was previously. Before, I just kind of took my precautions – I did my best to stay out and that kind of thing. But now, I find myself getting stressed out at the grocery store when people are too near me. And I find myself actively avoiding certain situations, even more so than I did before … So it’s just – I feel a lot more anxious, right now, when I’m out in public”* (BC, T2).

Such concerns were worsened by the rise of new strains of COVID-19, reigniting fear of uncertainty of the severity of the disease and risk of a greater spread: *“Everyone now is talking about the Delta variant and it’s just so mentally draining”* (ON, T3). These fears led to participants experiencing a greater sense of hopelessness due to concern that the current resources to combat COVID-19 would not be effective long-term:

> *“Just hearing that it is more deadly and more contagious, is definitely a worry. Because I know before people were thinking, well, you know, there are the vaccines coming out, so once I get it I’ll be fine. But you know, the findings that maybe it doesn’t work completely against the variants, that kind of makes it seem like there is no solution in place”* (ON, T2).

#### Perception of policy response

Participants had mixed reactions to government policy response in all three regions. In Canada, the Canada Emergency Response Benefit (CERB) program, which provided financial support to Canadians whose employment income was affected by COVID-19, was viewed overwhelmingly as having a positive impact by alleviating financial strain. For students in college and university, having the financial support allowed youth to focus on their education and limit additional stressors:

> *“I think the government definitely did help, especially with CERB cheques and things like that … having the government step in and financially assist me during this time was hugely beneficial, and allowing me to continue with my education, and not be stressed about money”* (BC, T1).

On the other hand, some participants in Ireland felt the COVID-19 Pandemic Unemployment Payments (PUP) were a source of stress, with some young person worrying *“that they’re going to make everyone pay back for COVID pay in tax”* (Ireland T3). Others felt frustrated by *“COVID pay”* (Ireland T3), causing tensions between those who received it and those who did not: *“My friend signed on to the COVID pandemic payment … I stopped talking to her for about four days, I was like … ‘I have to do 20 hours in three days. I am exhausted and you’re rubbing it in my face’”* (Ireland, T2).

While youth expressed contentment with restrictions implemented to keep people safe, a number of concerns were also raised. Over time, dissatisfaction centred around issues of changing guidelines and continuous lockdowns put in place by each region’s government: *“I think the way they’re going about it, as in a little-by-little, and then they open it a little bit and then they go back again, think that’s doing a lot of damage to people’s mental health”* (ON, T2).

#### Absent youth voice in policymaking

A growing concern raised across sites and timeframes was a consistent lack of youth voice. Youth did not see themselves reflected in policy or feel they had an impact to steer outcomes, though they noted the importance of their voice for realising change for the youth population. In terms of policy decisions, youth expressed discontent with *“no youth at that table”* (BC, T1) and felt *“brushed to the side”* (Ireland, T3), describing, *“I feel like we have no say and we’re just following what other people are telling us to do”* (ON, T2). Lack of attention to youth issues contributed to a sense of invisibility: *“It’s like we don’t exist”* (Ireland, T2).

Despite desire to be involved in government and decision-making processes, youth noted minimal opportunity to engage: *“To a person who is young, them being able to put their voices out, that ability is very much hampered down, just because of these systemic barriers that are happening, and forces that work against young people”* (BC, T1). The absence of youth participation was most apparent concerning decisions around education. Many felt dissatisfied with schools closing and re-opening:

> *“Being a student is unique, because I feel like most students were placed in a position where they didn’t really have a lot of power and we felt really passive in how we were receiving information and how decisions were being made in terms of our education*” (ON, T3).

As the last group to get the vaccine, youth raised concerns about returning to school: *“My university has mentioned that we would be opening up in September* [2021] *despite the fact basically no one would be up for vaccination until October at the earliest. I feel like that’s something someone could have pointed out”* (ON, T2). Lack of choice regarding decisions to continue school virtually or in-person created strife among university-aged students: *“Going back to campus full time without any online options, I don’t think that a young person would necessarily make that decision”* (BC, T3). Frustration continued to grow over time as the perception of neglect persisted: *“There’s been such a lack of concern for university age people, you know? That I think people are getting a bit disgruntled”* (Ireland, T3).

Youth recognized the value in having their opinion shared despite being young: *“Just because you’re 20 doesn’t mean your experience is worth less”* (BC, T3). Desire for involvement stemmed from an understanding of being future decision makers: *“We’ll be the next people in-charge of things. Every generation can build on the last generation … I think that’s what makes young people in the world so important, because we can all learn from people’s past mistakes and build on that and stuff”* (Ireland, T2). With this mindset, youth described the importance of engaging in activism and online petitions and voting when of age in order to effect change.

#### Unfair judgement of youth role in the pandemic

A recurring sentiment among participants was the perception that youth were being disproportionately blamed for the spread of infection in the media. Youth described feeling *“unfairly targeted”* (BC, T1) upon experiencing criticism for breaking rules and not taking the pandemic seriously:

> *“We’ve all been labelled like this, you know, ‘it’s the young spreading the virus’, ‘it’s the young that don’t care’, but it’s not like that, that’s not true, like I’m taking all this very serious. It’s affected my life in ways that I could never have even imagined”* (Ireland, T1).

In light of these accusations, youth countered such blame by pointing out how they are vulnerable to infection as the last age group to be eligible for the vaccine, as well as a demographic notably on the front lines of customer service and hospitality jobs, placing them at increased risk compared to those with the option to work from home:

> *“… [Young people] are sort of being blamed for a lot of the spread, but also, that age category is probably a lot of the people who’re working minimum-wage, front-line jobs, being forced to be out in places, taking public transit, living with roommates … And we’re going to be the last to be vaccinated which is, again, frustrating”* (ON, T2).

#### Vaccination in the face of hesitation

Overall, youth expressed willingness to get a COVID-19 vaccine. For those with concerns, hesitancy was raised surrounding uncertainty about the speed at which the vaccines were developed and rolled out in addition to not knowing the long-term impacts. Such concerns were most commonly raised at T1, prior to vaccines becoming available: *“I’m not anti-vaccine, but I feel like they are rushed … You never know what the side effects are in a year or ten years from that vaccine, right? That’s something that I worry about”* (BC, T1). Some youth felt like the misinformation online contributed to their uncertainty about the vaccine:

> *“I have mixed feelings about [the vaccine] … I feel like I feed in to Facebook been like, oh, it will make you have a brain damage … don’t take it, [you’ll] never have children and stuff like that. But I feel like if it was that bad, they wouldn’t be giving it to people. So, I just look at actual scientific side of it rather than feed into random people on Facebook”* (Ireland, T2).

To address these concerns, youth described needing more information, presented in a way that explains how the vaccine works:

> *“I want to know information and I want to learn about it. I wouldn’t let anyone put any vaccine into me without knowing about it. So, I think what has really worried me is that the information and the responses have been so, so vague to a point where it seems really shady”* (ON, T2).

Despite hesitation, youth discussed several motivating factors that led them to get a COVID-19 vaccine, primarily for functional reasons and as a means to return to normalcy by T2 and T3. Youth noted how a vaccine would provide a sense of safety: *“I am at ease that something is protecting me”* (ON, T3). More than just in a practical sense, vaccines gave participants a renewed sense of optimism:

> *“What is exciting about the vaccine is the potential to dream about the future again in a meaningful capacity. In a way, COVID took away the ability to conceive of any future because the future became indistinct, right. But having the vaccine allows you to go back to imagining a future that is with other people which is important”* (ON, T2).

### 5. Navigating COVID-19 information

#### Confusion around pandemic information presented by the government

Youth found pandemic information confusing and containing too much jargon: *“It was not easy to understand. Just a lot of big words and government words”* (BC, T1). As a result of inaccessible language, youth were deterred from seeking out information: *“I don’t listen to it anymore because it’s too confusing, it’s too much information being thrown out in 5-minute speeches”* (Ireland, T1). Over time, as guidelines changed, participants felt it was hard to follow and keep up to date with current guidelines: *“I think maybe the government could’ve done a bit better of mitigating some of that ambiguity and uncertainty, and helping people cope with the ever-changing guidelines and protocols”* (BC, T1). The need for youth-friendly information was addressed:

> *“I feel like they should have like some younger people on the government’s team just to like explain what some things mean to the younger generations … me and some of my friends have read some stuff from the government website, and we’re just like, ‘what does even half of this mean?’”* (ON, T1).

#### Accessing COVID-19 information through multiple forms of media

The primary sources where youth accessed pandemic information were the government and news media’s social media, webpages, and daily addresses. Having options tailored to the needs of the individual was beneficial:

> *“I really like the Prime Minister’s address … because I have dyslexia, I don’t really like reading, I don’t like going through long articles. I need to just hear it … So, I just appreciated someone putting everything into a concise press conference so I don’t have to go hunting for all the articles, hunting for the information and everything”* (ON, T1).

Social media was commonly discussed, as participants described using various platforms such as verified news outlets and government run Instagram, TikTok, Twitter, Reddit, and YouTube to access information from people or organizations whom they trust:

> *“I think the government actually did a decent job of putting things on Instagram. I wasn’t really active on any other platform, but just seeing like, you know, the infographics about, here’s what you’re allowed to do”* (BC, T3).

Youth were astutely aware of the need to find reliable sources and wade through misinformation, citing the spread of false information as a barrier to staying informed:

> *“It’s very hard to avoid any kind of misinformation or just complete lies about it. That’s kind of a difficult thing, and I’d say that’s where a lot of difficulty in miscommunication comes from is just because people’s primary new source is the internet, which is not always very reliable”* (Ireland, T3).

### 6. Transitioning to life after the pandemic

#### Cautious readjustment to a new normal

Young people anticipated challenges with re-adjusting to life after the pandemic with years of diminished social interaction and physical distancing, particularly felt in the midst of lockdowns: *“Everyone’s afraid of each other”* (Ireland, T2). Participants described a persistent cautious outlook when attending public events or gatherings, with one participant saying, *“I think immediately after the pandemic, there’s going to be a lot of people who are a little skittish about re-integration into crowds and restaurants”* (ON, T2). Such challenges with reintegration were recognized *“as almost a trauma response, where if someone gets too close to you, you feel unsafe”* (BC, T1).

Some participants grew comfortable with spending time alone, with long-lasting impacts on their desire for socialization: *“I’ve gotten used to my own company and I’m starting to enjoy just being with myself but that’s also a negative”* (Ireland, T2). Participants recognized the importance of social interaction, though felt as though transitioning from spending time alone to engaging with others in person might be challenging: *“I feel like it’s going to be hard to go back to real life, because I’m so used to not leaving my house, now, and I’m kind of like a bear in hibernation, now. I don’t know if that’s going to be an easy transition”* (BC, T1). Balancing the needs for social interaction with the comfort of spending time alone was thought to be a difficult balance to navigate, with concerns for long-term mental health impacts. Participants recognized these challenges as collective trauma: *“Some people will be traumatized by it and may never heal from it”* (ON, T2). Without adequate supports in place, youth feared for their ability to heal moving forward.

#### Concerns for the future

When considering their futures in a post-pandemic world, participants expressed concern for the state of the economy as well as their own job prospects: *“I think economically Ireland will have a long time recovering and especially for when I’m older and I’m buying a house. I feel like the prices will still be really high”* (Ireland, T1). In light of these concerns, several youth noted that they re-evaluated their plans and goals for the future to adjust to a new world and changes to their own self, considering, *“What is post-COVID for me now, because I am in a much different place than I thought I would be”* (ON, T3). In light of these fears, youth described the need for support, though had concerns that their needs would not be met, particularly for those without a strong social support network:

> *“I do think [young people] face unique circumstances and I think because of a lot of income inequality, like the general expectations for new graduates, I think young people have a lot of challenges that have been exacerbated by the pandemic, and I feel like if there aren’t the correct social safety nets in place. It can leave a lot of young people behind … You know, not everyone has a family that they can rely upon during these sorts of times, and I think we just have to make sure that we’re looking out for everyone”* (BC, T1).

#### Optimism about positive social changes

Despite hardships endured during the pandemic, youth were able to identify silver linings and potentially positive long-term changes in terms of the workplace, education, hygienic practices, and social justice. Youth hoped to see flexibility regarding work-from-home policies, *“where workplaces are more lenient, [and] people understand things like childcare”* (BC, T1), with improved accommodations for employee needs*: “I think more people are going to be opting to work from home if they’re allowed to because commuting to work doesn’t really do much for them”* (ON, T2).

Youth were optimistic about improved health beyond the pandemic. Continued hygienic practices to prevent spread of illness beyond COVID-19 were discussed in a positive light: *“Sanitizing, washing our hands, wearing masks when we’re sick, taking time off to stay at home when we’re sick, I feel like those are going to have long-term affects within the workplace, and within school”* (BC, T2). While exacerbated mental health challenges were noted as a continued uphill battle to address, the pandemic sparked more conversation around mental health, trauma, and help-seeking that resulted in de-stigmatization, which was thought to be a step in the right direction:

> *“[The pandemic] really highlighted the impact that social and global events can have on someone’s mental health, and so I feel like I saw a lot of people kind of advocating, either trying to point out programs or places that people could get help”* (ON, T3).
>
> *“I hope in the recovery of Ireland and the economy of Ireland and stuff, they don’t forget about mental health because it was such an important thing during COVID and I hope that’s, that slowly doesn’t fade away” (Ireland, T1)*.

Because of the political discourse throughout the pandemic with attention to social justice issues, youth were optimistic that some of the structural inequities brought to light would be addressed in society: *“I think that will be the big frontier, because I think the pandemic, for all the bad, does provide an opportunity to rectify some of the inequalities we’ve seen exposed through it”* (BC, T1). Inherently tied to these changes was a developed sense of compassion and empathy among youth and others, and the belief that change is possible:

> *“I’m hoping this period helps develop empathy and kindness in people … I’m hoping that this is an opportunity for people to kind of reflect on the skills that they offer in that sense and hopefully grow in that sense, and using those newfound skills and newfound parts of themselves to engage with their communities and to take care of their communities, whether that’s physically or by leading it on a kind of social change front or whatever that may look like”* (ON, T1).

In this sense, youth expressed actionable strategies to use their voice through social media and partaking in social justice movements to pressure policymakers: “*We deserve health and equity, and we have the resources and the capability to have those things, we just need to make it happen”* (ON, T3). Participants found hope in this belief, with the change starting within themselves: *“I know me, for one, I’ve come out of it a better person”* (Ireland, T2).

### 7. Youth-led recommendations for government and service response

Four subcategories about key recommendations for government and service response were generated from the data, which related to strategies for amplifying youth voice, communicating public health information effectively to youth and combatting misinformation, optimizing services post-pandemic, and planning for future pandemics.

#### Youth voice needed in decision making

Participants raised the importance of incorporating youth voices in policy decisions, particularly when directly impacted by the outcome. Youth articulated the need for their voices to shine through research so that policymakers understand their needs, with attention to questions such as, *“How are you living your life? What are your priorities? What are your concerns?”* (ON, T3). School was recognized as the best way to reach a broad spectrum of youth: *“I feel like the only way you could really collect information from a bunch of young people is through school”* (ON, T3).

In addition to research, youth expressed desire for more opportunities for leadership to develop skills in advocacy and bridge the gap between youth and those in charge. Adults in positions of power have an important role in creating such opportunities, where leaders *“who have paved the way for other youth to use their voice [are] really important”* (BC, T1). Opportunities at school for youth to address issues were discussed: *“I feel like there could be maybe like a group of students and young people who can kind of just voice the opinions of everyone on their behalf and stuff and maybe just offer up suggestions”* (Ireland, T1). On a grander scale, the implementation of a youth advocate role in government was mentioned to communicate needs *“instead of someone just kind of guessing for young people”* (Ireland, T1).

The importance of diversity in youth experiences was recognized, with understanding that historically, persistently, or systemically marginalized youth with important stories to share may be ostracized from participation in research and decision making:

> *“I feel like there are definitely people who should be involved but the people who should be involved are not usually the people who can be involved. Like if you are poor and in an immigrant household you are probably at home taking care of you siblings and have no time to sit on committee meetings”* (ON, T2).

#### Communicating public health information effectively to youth and combatting misinformation

Youth discussed the need to continue to *“use social media in a way that you promote responsible information or reliable information”* (ON, T3) because *“that’s where a lot of young people spend a lot of their time”* (BC, T3). Concise, digestible content in the form of infographics was said to be an effective way to disseminate information, promote services, and encourage service uptake: *“None of the BS, you know … cut all the medical jargon out … explain it simply”* (Ireland, T1).

With regard to sharing COVID-19 information, youth thought it *“would be helpful if there was more clarity as to why each restriction was put in place”* (Ireland, T1) to combat frustrations with fluctuating and sometimes contradictory restrictions and to improve adherence to guidelines.

“I think it would be helpful if there was more kind of like clarity as to why each restriction is put in place, because people would be like ‘Hang on. So we can’t do this, but people can like, come into the country and leave the country.’ And like ‘We can do this at school, but we can’t do this in shops’. And they’ll be like, why is that? And they kind of get frustrated and be like, I’m not listening to them anymore. So I think it would be helpful if there is more thorough explanations as to why, like maybe you can do this, but not that, and stuff.” [el]

Aiming information at parents is another way to spark dialogue between parents and their children about mental health: *“I think by targeting adults, that would help the situation for younger people to feel more comfortable talking about their issues”* (BC, T3).

Aside from social media, school was reiterated as a means to convey information to youth, teach emotional literacy, and provide resources: *“I think getting information to students through institutions, I think it’s a very smart move. Like keeping up with your school’s newsletter”* (ON, T3).

Creating visibility surrounding resources and mental health can promote help-seeking, as recognized by a university student in BC: *“I feel like sometimes it’s like you have to go out and seek it, but it’d be nice if universities sent out emails midway through the semester, just to tell people about that”* (BC, T2).

A key issue raised by youth regarding accessibility of virtual information was the prevalence of misinformation and knowing how to identify reliable sources. As a generation who grew up with social media, the need for strategies to address these issues was apparent. Youth expressed desire to learn about internet literacy and critical thinking skills in the classroom:

> *“I think internet literacy, where if people grow up and they learn, OK, not everything on the internet is true … If that thought process was taught in school, then it wouldn’t really be as big a problem as it is today”* (ON, T2).

Similarly, accountability within the platforms themselves and in government was suggested as a welcomed solution:

> *“I think holding the people accountable that spread misinformation will help and there being less bots or problematic ideas going … tech companies holding people accountable, or governments holding people accountable”* (ON, T3).
>
> *“There’s a lot of false information that spreads very quickly and there definitely needs to a little bit more concern on the platforms part to ensure that stuff like that does not happen”* (ON, T3).

#### Post-pandemic services: enhancing service delivery to improve outcomes

An issue brought to light during the pandemic was the lack of affordable and accessible mental health services, worsened by intensifying mental health problems experienced during this time. For youth, the pandemic presented an opportunity to address some of the issues faced within the system that impact youth specifically. Despite a desire to seek help, lack of affordable and timely services created a major barrier to care:

> *“A lot of people want mental health services and either they can’t afford it or they’re on like a six-month, one-year waiting list”* (ON, T3).

To address this issue, youth recommended expanding social safety nets to cater to the needs of this population:

> *“Starting with the expansion of disability benefits, making it so that people who are on disability are not living in poverty, that would be a good start, because we are going to have a fall out in terms of the people who have been affected by COVID”* (ON, T3).

In the aftermath of COVID-19, there was an expressed desire for priority to be given to mental health services to counterbalance the mental health challenges that were worsened by increased stress and decreased access to services:

> *“The government’s definitely going to have to implement some sort of strategy, like for aftercare, I feel. I just feel like it’s going to be a domino effect and it’s going to go on for years to come … You just don’t know what kind of effect that would have on somebody”* (Ireland, T2).

Post-pandemic services should also continue to offer virtual care according to participants. While many youth declared a preference for in-person services, there was recognition of the value in having options:

> *“[Services should] still be definitely available online, especially because some people, you know, maybe aren’t comfortable [with] the face-to-face environment yet, or maybe are immunocompromised or just physically can’t get there”* (BC, T3).

#### Planning for future pandemics

In a future pandemic, youth emphasized that personalizing messaging to all demographics is important. In the youth context, this can be seen as communicating information regarding an infectious disease in a way that is easy to understand, digestible, and does not instill fear:

> *“I really like simplicity, like getting to the point. Easy facts. Anything factual information that has key points and bolded and have diagrams, pictures, or even, like, an example, analogies, et cetera, would be really important to understand”* (BC, T1).

Ensuring resources and funding are available would help youth feel supported in times of crisis. Not all youth have strong familial relationships to provide safety and protection, requiring implementation of safety nets:

> *“Far too often, the young-person demographic, so like not a minor, but not a middle-aged person, is often ignored in planning stages. It’s assumed that we can operate as those people have, but I think we have a much less robust support system, and even just crisis funds and things like that. I think that needs to be accounted for”* (ON, T2).

In light of these recommendations, it is evident that the pandemic is not simply an acute public health crisis but rather a historical event that will have lasting impacts on youth health and wellness. COVID-19 exposed gaps in service delivery for this population, where ongoing needs remain to be comprehensively addressed.

## Discussion

This study explored youth experiences during COVID-19 over time and offered youth-derived recommendations for government and service responses to pandemics and other public health emergencies. Our longitudinal qualitative findings yielded key trends indicating the ways in which COVID-19 impacted participants at multiple time points over the evolving pandemic, with consistent findings across regions (BC, Canada; Ontario, Canada; Ireland). Youth described their experiences of coping with the many hardships of the pandemic and adapting to new ways of accessing services, but they also identified areas for personal growth through the hardship. Their views and attitudes about the pandemic were mixed, and they found it challenging to navigate the complex information provided about the pandemic. Challenges also arose with transitioning to a post-pandemic life. Youth leveraged their experiences to provide recommendations for responses by governments and service-providing organizations for future pandemics and public health emergencies.

Findings were consistent at each time point, despite some differences in the cohorts across the three sites. This may be due to the similar COVID-19 responses in these regions, including lockdown measures, vaccination rollout, testing, and contact tracing [41]. Their adaptive responses all included a series of lockdowns comprising of stay-at-home orders, school and non-essential business closures, a vaccination strategy that prioritized vulnerable populations and essential workers, mandatory use of face coverings, and providing income support to affected workers [41]. Furthermore, ON, BC, and Ireland have integrated youth services that provided virtual mental health services to youth throughout the pandemic [30,35,42]. While Ireland to date has more confirmed COVID-19 cases per capita than ON and BC, this did not seem to influence the impacts on youth [43]. These findings align with observations in similar populations in other high-income countries with integrated youth services [41]. Nevertheless, it is crucial to acknowledge that further research on the impacts of the COVID-19 pandemic on youth in areas with lower income, a varied COVID-19 response, and/or without integrated youth services would be beneficial to assess the applicability of these findings across wider regions.

The challenges experienced in response to the COVID-19 pandemic were numerous and reflect the emerging literature. For example, the loss of key experiences has been identified as a factor affecting young people in important ways during the pandemic [44,45]. Educational challenges presented in the form of difficulties adjusting to online learning [24,46–48]. Job loss and financial strain contributed to stress over time due to sudden economic instability and future uncertainty [49,50]. Essential workers struggled with a return to risky work environments, facing the stress of contracting COVID-19 as well as managing new roles and responsibilities for enforcing public health guidelines [51]. An additional hardship noted was the increased prevalence of racism and xenophobia. Anti-Asian attitudes in the age of COVID-19 have been documented in the American context [52–54], noting higher rates of harassment and health disparities throughout the pandemic due to harmful rhetoric regarding COVID-19 origins. Difficult-to-follow public health information was an additional challenge, as was initial vaccine hesitancy [55–60], although attitudes shifted toward favouring vaccines with time. With the myriad of challenges came a decline in mental health and mixed effects on substance use behaviors, particularly noted in T1 and T2, as is documented in the literature cross sectionally and longitudinally [16–20,29,61–63].

With the complex changes in contexts, environments, and lifestyle, youth also experienced disruptions in health service accessibility, including mental health service access. These disruptions may have further constituted a barrier to health and wellness [31,64], although acceptance of virtual services seemed to increase with time. Isolation, financial strain, and setbacks in educational and professional development in the context of reduced service accessibility may be contextual factors that led to the short-term impacts on mental health; this trajectory could also extend into long-term impacts on mental health and substance use, which is an important area for ongoing research as the pandemic resolves [65].

Despite the hardships experienced, youth also identified several strengths and opportunities for growth during the COVID-19 pandemic. Youth described picking up new skills and hobbies such as cooking, exercise, and creative endeavors, which helped with overall wellness. In some cases, mental health improved due to reduced demands on time and increased time to self-reflect, slow down, and recover from burnout. In this sense, the pandemic was a catalyst for change where help-seeking and engagement in services were realized for the first time. Access to green spaces was a positive factor; indeed, this has been shown to reduce symptoms of depression, anxiety, and stress [66–68]. A small body of research discusses the positive developmental opportunities, strengths, and effective coping strategies leveraged by youth during the changes in the pandemic [8,69]. Another body of research outlines post-traumatic growth in various populations during this time [69,70]. Together, these findings can serve to identify strengths, protective factors, and resiliencies that can be harnessed to support mental health and wellbeing during challenging periods in history, from a pragmatic and strengths-based lens. Indeed, youth remained optimistic about the potential for the pandemic to act as a catalyst for positive social change.

Youth noted a lack of attention to youth needs, consideration of unique harms experienced by youth, and youth engagement and leadership in developing solutions throughout the pandemic response. To this end, consistent with extant literature [71,72], continued efforts should be made to include youth in research and policymaking by centering youth issues and engaging with youth as equal stakeholders/rightsholders. A report by the Children and Youth in Challenging Contexts (CYCC) Network (2013) offers best practices for engaging with youth to address social exclusion and mitigate power imbalances by highlighting the importance of collaboration [73]. Organizations, schools, and all levels of government should enhance capacity for creating opportunities where youth can speak on relevant issues and be heard immediately. The creation of student/youth advocacy groups and the development of government liaison positions can help to bridge gaps and encourage political participation.

Based on the current findings and the emerging literature, examples of solutions that youth might propose to better address future pandemics and public health emergencies include the use of verified, fact-checked social media posts to clearly communicate public health information in a consistent manner [74,75], together with strategies to eliminate help-seeking barriers and support system navigation [76,77]. Incorporating age-appropriate internet literacy into school curricula to help cultivate critical thinking skills and discern factual information from unreliable sources was also discussed. Free or affordable mental health services are needed, as well as attention to accessibility needs. Attention should be given to enhancing access for those with minimal technological literacy skills [78]. Financial supports and means to safely socialize, exercise, and alleviate symptoms of mental illness are additional youth priorities. In a similar vein, resources should be distributed to address safety concerns for those in unsafe households, who may be at risk of emotional or physical abuse during periods of lockdown. Additional novel goals and solutions may be proposed through ongoing and appropriate youth engagement in policy, service, and research design.

### Strengths and limitations

This study provided longitudinal insights that capture a substantial period of change during the evolving COVID-19 pandemic. Spanning multiple jurisdictions with large samples, the study identified cross-cutting experiences reported by a wide variety of youth. With youth engagement throughout, the research questions and approaches were informed by youth voices. However, limitations of this study include varied sampling strategies and timelines across study sites, complicating comparisons between sites, as well as attrition. While flexibility in sampling strategies is a strength of qualitative longitudinal research design for reasons including enhancing generalizability of findings, comparability can also be a limitation. The variability in recruitment approaches across different sites may have hindered the ability to discern consistent patterns and trends over time. Given the longitudinal nature of this study, participant numbers declined over time. As the findings suggest, “COVID fatigue” may have contributed to study drop-out. Those who chose to remain in the study may differ from those who did not complete all three timelines. The use of virtual means to recruit and conduct the interviews expanded the geographical reach of the study, but limited it to youth with online access.

## Conclusion

This is the first known international longitudinal qualitative study to describe youth experiences during the COVID-19 pandemic across two Canadian jurisdictions and Ireland, with implications for policy and practice. Despite the many challenges encountered over the course of the pandemic, youth reported aspects of growth. For this population, pre-existing stressors were compounded by pandemic-related challenges and reduced access to care, signifying the importance of prioritizing youth mental health moving forward. Areas of need highlighted throughout the pandemic have implications for future policy decisions, with an opportunity to enhance health and social service delivery. Investment in resources rooted in youth-identified needs and priorities is vital to mitigate the long-term challenges associated with pandemic trauma among youth.

## Data Availability

Data cannot be shared publicly because of they are under the care of three guiding institutions. Opportunities to explore data access from the University of British Columbia should be made directly through Dr. Skye Barbic. For Ontario data, please connect with Dr. JL Henderson. For Irish data, please connect with Aileen O'Reilly.

## Acknowledgements

We are grateful to have conducted our research on the ancestral lands of many different Indigenous Nations and Peoples across what we now call British Columbia and Ontario. We would like to thank the youth participants, advisors, and research staff, who contributed valuable insights and shared their experiences throughout the duration of this project. We would also like to acknowledge with gratitude the research teams at Jigsaw, the Centre for Addictions and Mental Health, Foundry, and the University of British Columbia.

## Appendix A. Semi-structured interview guides

### T1 Interview Guide

1. How would you describe your life before COVID-19 started?
  a. Were you in school? Working? How much/often
  b. How was your social life?
  c. Hobbies?/activities
  d. Day in the life?
2. How would you describe your life now, in COVID-19 times?
3. As a young person, how has COVID-19 affected your everyday life?
  a. Has COVID-19 had any positive impacts on your life? What kinds of impacts?
  b. Has COVID-19 had any negative impacts on your life? What kinds of impacts?
4. What would you be doing different in your life now if it weren’t for COVID-19?
5. Many services for youth have changed in response to the COVID-19 pandemic.
  a. What were you accessing before COVID and what has changed for you?
  b. How could the changes have been better?
  c. Have you used virtual services? Why or why not have you accessed? What kind of services? How have they been working? What kind of remote technology have you used? (phone, text, video).
6. How hopeful are you feeling about Canada’s recovery from COVID-19?
7. What activities are meaningful for you or other youth to engage in to make a difference during the COVID-19 crisis?
8. What kinds of informational resources could you or other youth benefit from during the COVID-19 crisis?
9. How do you see your post-pandemic life?
10. As a young person, you may be of thinking about your future, for example school, career path, etc.
  a. How has your planning process changed due to COVID-19?
  b. How have your plans changed?
11. Imagine your life two years from now. How do you think COVID-19 will have impacted your life in five years?
12. Imagine your life five years from now. How do you think COVID-19 will have impacted your life in ten years?
13. How do you think youth see the pandemic and how seriously they are taking things?
14. What positive contributions are youth making to the pandemic situation?
15. Do you think young people can play a role in planning the response to this pandemic and future pandemics? What kind of role? What would this look like?
16. What role do you think youth are playing in decision-making (policy making, service planning) and how has that impacted the pandemic?
17. If you could make any suggestion to improve the pandemic response for youth, what would that be?
  a. Did the government response help you directly? Was the information easy to understand?

### T2 Interview Guide

1. How would you describe your life since the last time you were interviewed? Has anything changed for you?
  a. During the last interview, you were asked about school/college, or if you are employed… how has that changed for you, if at all?
  b. During the last interview, you were asked about your current social life… [you were missing friends, you have maintained an average social life, etc]… has that changed? How?
  c. During your last interview, you were asked about your hobbies or activities you enjoy and may not be able to do. Has that changed for you? How?
  d. Are you living in a rural area, town, or city? [Base next part off of their answer] Do you think living *[rural/in a town/city/village]* made your experience of COVID any different?
2. Since your last interview, as your experiences of life during the COVID-19 pandemic changed in any other way?
  a. Has COVID-19 had any positive impacts on your life since the summer? What kinds of impacts?
  b. Has COVID-19 had any negative impacts on your life since the summer? What kinds of impacts?
3. During your last interview, you were asked what you would be doing differently in your life if it weren’t for COVID-19. What (or what else) would you be doing now if it weren’t for COVID-19?
4. During your last interview, you were asked about services you may have used before/during COVID, and what it’s been like to use them.
  a. Have you been using services during COVID-19? Have you used mental health services, specifically?
  b. Since your last interview, have you continued using this service, or others, on a regular basis? Did you start or stop using any other services since your last interview?
  c. Since your last interview, have there been any additional changes to this service? What are your thoughts on those?
  d. *If relevant,* how was finishing up with that service during COVID for you?
  e. *If relevant and using a new service*
    i. Why did you decide to use these services?
    ii. What kind of support were you offered? (e.g., phone, video, F2F)
    iii. What did you find most helpful about these services?
    iv. What did you find least helpful?
    v. What were your expectations of the service? Were they met?
    vi. Were you satisfied with the services/would you recommend to a friend?
  f. Is there anything about that service that you didn’t mention the last time that you would like to talk about?
5. Something we are interested in is how mental health services have changed during the pandemic, including having more experience and capacity in providing online services.
  a. Do you think online services like this should continue to be offered after the pandemic?
  b. What would this look like when in-person services resume?
  c. Over the course of the pandemic, has your opinion about online services changed?
6. Do you have any more ideas about how service providers could make it easier for young people to engage with mental health services?
7. During your last interview, you might remember that you were asked about Canada’s recovery from COVID-19. How hopeful do you feel about that now?
  a. Compared to how you felt during your last interview, why are you feeling more hopeful/less hopeful now?
  b. What would make you feel more hopeful? Less hopeful?
8. During your last interview, there may not have been a vaccine available for COVID-19. Now vaccines are slowly being rolled out across Canada.
  a. How do you feel about the vaccine?
  b. Where have you been getting information about the vaccine?
  c. Have you gotten it, or do you plan to get it? Why/why not?
  d. What has influenced your opinion about the vaccine?
    i. What might make you excited to get the vaccine?
    ii. What might make you hesitant about the vaccine?
  e. How is this affected by your [identity factor]
  f. What about people in your family and social circles? What are they saying about the vaccine? How is this influencing your perspectives?
  g. What kinds of informational materials do youth need to make good decisions about the vaccine?
  h. Do you have any other thoughts to share with us about the vaccine?
9. During your last interview, you were asked about your life after COVID-19. How do you see your post-pandemic life now?
  a. Since your last interview, has anything about your plans for after COVID-19?
  b. Do you see COVID-19 having lasting impacts in our society? If yes, what would this look like? How would these affect your everyday life?
  c. You were asked before, but just in case anything has changed… Imagine your life five years from now. How do you think COVID-19 will have impacted your life in five years?
  d. Imagine your life ten years from now. How do you think COVID-19 will have impacted your life in ten years?
10. Finally, you might remember during your last interview, you were asked about how you think young people see the pandemic. Since then, do you think anything has changed about how young people see the pandemic and how seriously they’re taking things?
  a. What contributions are young people making now? Negative/positive?
  b. Do you think there is anything different about the role young people are playing in decision-making (policy making, service planning)?
  c. Is there anything else about how young people get information about COVID- 19 that you think would be helpful to say?
  d. What would you say now about the role young people play in planning a response to a pandemic?
  e. If you could make any suggestion to improve the pandemic response for young people, what would that be?
11. Is there anything else we haven’t covered that you would like to add or think we would be interested?
12. Do you have any questions?

### T3 Interview Guide

1. How would you describe your life since the last time we spoke? Has anything changed for you?
  a. The last time we spoke you told me you were in school/college/working/unemployed…how has that changed for you, if at all?
  b. Last we spoke you told me [you were missing friends/had an average social life, etc]…has that changed? How?
  c. Last time we spoke, you told me you enjoyed/weren’t able to do [hobbies/activities]…has that changed for you? How?
  d. Are you still living in a rural area, town or city? Do you think living *[rural/in a town/city/village]* made your experience of COVID any different? **NB. If young person disclosed any relevant identity factor, interviewer will explore how this might have affected their experience throughout the interview**
2. Has your experiences of life during the COVID-19 pandemic changed in any other way since the last time we spoke?
  a. Has COVID-19 had any positive impacts on your life since the we last spoke? What kinds of impacts?
  b. Has COVID-19 had any negative impacts on your life since the we last spoke? What kinds of impacts?
  c. We’ve heard from some young people that grieving has been different during the pandemic, for example grieving for the loss of a loved one. Have you been affected by grief during the pandemic? How has this gone? How has it been different due to the pandemic?
  d. Last time we spoke, Covid-19 rates were quite high in [insert country]. How do you think people with Covid-19 are perceived? Do you think where people are from (I.e. city or countryside) influences their opinion Covid-19?
3. How would you describe your mental health since we last spoke?
  a. General probes: (If it improved, what factors led to that improvement? If it declined, what lead to the decline?)
4. How would you describe your substance use since we last spoke?
  a. General probes: (How has it changed? If it increased, what factors led to that improvement? If it declined, what lead to the decline?)
    i. Remember to probe based on the substance (e.g. cocaine use may decrease, but cannabis use may increase, etc.)
5. There has been a lot of discussion about mental health during COVID-19. How do you think this has affected people’s attitudes about mental health?
  a. General probes: (How have they changed? If improved, what factors led to that improvement? If worse, what lead to the decline?)
    i. Probe stigma
6. The last time we spoke, we talked about what you would be doing differently in your life if it weren’t for COVID-19. What (or what else) would you be doing now if it weren’t for COVID-19?
7. The last time we spoke, we talked about how services for young people have changed in response to the COVID-19 pandemic. (At the time, you told me about what it was like to use [previously mentioned service], before/during COVID…)
  a. Have you continued using this service/started using this service again/finished engaging with this service since we last spoke?
  b. Have there been any additional changes to this service since we last spoke? What are your thoughts on those?
  c. Some new services have been created during COVID-19. Have you heard of any new services in your community? What kind of services (e.g., virtual, in person, specific cultural groups, etc.)? How has this affected your access to services?
  d. *If relevant,* how was finishing up with that service during COVID for you?
  e. *If relevant and using a new service*
    i. Why did you decide to use these services?
    ii. What kind of support were you offered? (e.g., phone, video, F2F)
    iii. What did you find most helpful about these services?
    iv. What did you find least helpful?
    v. What were your expectations of the service? Were they met?
    vi. Were you satisfied with the services/would you recommend to a friend?
  f. *If relevant and using any MH service:*
  i. How did the restrictions affect your ability to benefit from the services? For example, if you were learning new skills in therapy, were you able to apply them when we were under restrictions? How might your experience with this have been different due to COVID- 19?
  g. Is there anything about that service that we didn’t chat about the last time that you would like to tell me about?
8. The last time we spoke, we talked about how many mental health services now have more experience and capacity to provide online services and whether you thought online services should continue to be available after the pandemic. Has your opinion about this changed at all?
9. Do you have any more ideas about how we could make it easier for young people to engage with mental health services?
10. In addition to using services virtually, many young people are using technology more than they were before the pandemic: personally, at work, in services and in other areas of life. How is this impacting you? Do you think this is a positive or a negative change? Why?
11. You might remember that we talked about Ireland/Canada’s recovery from COVID- 19 the last time we spoke. How hopeful do you feel about that now?
  a. Why are you feeling more hopeful/less hopeful now that you were when we last spoke?
  b. What would make you feel more hopeful? Less hopeful?
12. Since the last time we talked, there have been changes in how available COVID-19 vaccines are and in how many people have received them..
  a. Have you gotten it, or do you plan to get it? Why/why not?
  b. How have your thoughts on the vaccine changed? What caused them to change?
  c. What about people in your family and social circles? Have their perspectives on the vaccine changed? How is this influencing your perspectives?
  d. Do you have any other thoughts to share with us about the vaccine?
13. The last time we met, we talked about your life after COVID-19. How do you see your post-pandemic life now?
14. Has anything about your plans for after COVID-19 changed since the last time we spoke?
15. As society opens back up, we’ll all be returning to in-person activities soon. How do you feel about this? Is there anything about this that makes you feel positive or hopeful? Is there anything about this that makes you feel afraid or nervous?
16. Do you see COVID-19 having lasting impacts in our society? If yes, what would this look like? How would these affect your everyday life?
17. I asked you this question before, but I will ask it again… Imagine your life five years from now. How do you think COVID-19 will have impacted your life in five years?
18. Imagine your life ten years from now. How do you think COVID-19 will have impacted your life in ten years?
19. Finally, you might remember the last time we met we talked about how young people see the pandemic. Do you think anything has changed about how young people see the pandemic and how seriously are they taking things?
  a. What contributions are young people making now? Negative/positive?
  b. Do you think there is anything different about the role young people are playing in decision-making (policy making, service planning)?
  c. What would you say now about the role young people play in planning a response to a pandemic?
  d. If you could make any suggestion to improve the pandemic response for young people, what would that be?
  e. Is there anything else about how young people get information about COVID-19 that you think would be helpful to say?
20. Is there anything else we haven’t covered that you would like to add or think we should know?
21. Do you have any questions?

